# Radiological Imaging of Viral Pneumonia Cases Identified Before the COVİD-19 Pandemic Period and COVİD-19 Pneumonia Cases Comparison of Characteristics

**DOI:** 10.1101/2022.05.11.22274305

**Authors:** Rana Günöz Cömert, Eda Cingöz, Sevim Meşe, Görkem Durak, Atadan Tunacı, Ali Ağaçfidan, Mustafa Önel, Şükrü Mehmet Ertürk

## Abstract

**Background:** Thoracic CT imaging is widely used as a diagnostic method in the diagnosis of COVID-19 pneumonia. Radiological differential diagnosis and isolation of other viral agents causing pneumonia in patients gained importance, especially during the pandemic period.

**Aims:** We aimed to investigate whether there is a difference between the CT imaging findings characteristically defined in COVID-19 pneumonia and the findings detected in pneumonia due to other viral agents, and which finding may be more effective in the diagnosis.

**Study Design:** The study included 249 adult patients with pneumonia found in thorax CT examination and positive COVID-19 RT-PCR test and 94 patients diagnosed with non-COVID pneumonia (viral PCR positive, no bacterial/fungal agents were detected in other cultures) from the last 5 years before the pandemic. It was retrospectively analyzed using the PACS System. CT findings were evaluated by two radiologists with 5 and 20 years of experience who did not know to which group the patient belonged, and it was decided by consensus.

**Methods:** Demographic data (age, gender, known chronic disease) and CT imaging findings (percentage of involvement, number of lesions, distribution preference, dominant pattern, ground-glass opacity distribution pattern, nodule, tree in bud sign, interstitial changes, crazy paving sign, reversed halo sign, vacuolar sign, halo sign, vascular enlargement, linear opacities, traction bronchiectasis, peribronchial wall thickness, air trapping, pleural retraction, pleural effusion, pericardial effusion, cavitation, mediastinal/hilar lymphadenopathy, dominant lesion size, consolidation, subpleural curvilinear opacities, air bronchogram, pleural thickening) of the patients were evaluated. CT findings were also evaluated with the RSNA consensus guideline and the CORADS scoring system. Data were divided into two main groups as non-COVID-19 and COVID-19 pneumonia and compared statistically with chi-square tests and multiple regression analysis of independent variables.

**Results:** Two main groups; RSNA and CORADS classification, percentage of involvement, number of lesions, distribution preference, dominant pattern, nodule, tree in bud, interstitial changes, crazy paving, reverse halo vascular enlargement, peribronchial wall thickness, air trapping, pleural retraction, pleural/pericardial effusion, cavitation and mediastinal/hilar lymphadenopathy were compared, significant differences were found between the groups (p < 0.01). Multiple linear regression analysis of independent variables found a significant effect of reverse halo sign (β = 0.097, p <0.05) and pleural effusion (β = 10.631, p <0.05) on COVID-19 pneumonia.

**Conclusion:** Presence of reverse halo and absence of pleural effusion was found to be efficient in the diagnosis of COVID-19 pneumonia.

## Introduction

Viruses are the most common cause of respiratory tract infections. It has been reported that viruses such as influenza, HPIV, Adenovirus, RSV, HMPV can cause lower respiratory tract infections in individuals with both normal immune systems and immunodeficiency; It is known that viruses such as rhinovirus, endemic coronaviruses, CMV, Herpes Simplex Virus (HSV), Varicella Zoster Virus (VZV), HBoV can cause lower respiratory tract infection only in those with immunodeficiency.^1^

Coronavirus disease (COVID) was first reported by the World Health Organization (WHO) on December 31, 2019, with pneumonia cases of unknown origin being reported in Wuhan, China, and then reached the pandemic stage in March 2020.^2^

SARS-CoV-2, the causative agent of COVID-19 disease, is an enveloped virus whose genetic material consists of single-stranded RNA. The RT-PCR test, in which viral nucleic acid is detected, is accepted as the gold standard for the detection of SARS-CoV-2 virus.^3^

It is reported that COVID-19 infection can be examined in 3 stages, including the first asymptomatic period, secondly the upper and lower respiratory tract response, and then widespread lung involvement that can progress to ARDS.^4^ In the COVID-19 disease, approximately 80% of the patients are asymptomatic or limited to mild to moderate symptoms in the first two stages; It is reported that in the remaining 15-20% of the patients, pulmonary ground glass opacity-consolidation is detected as a radiological finding due to the inflammatory response in the lung.^4^

If there is no risk factor for the progression of the disease in patients with mild clinical symptoms suspicious for COVID-19, there is no imaging indication, and imaging should be performed in cases with worsening respiratory system symptoms; It has been reported that imaging can be performed to provide medical triage in cases with high suspicion for COVID-19 with moderate-to-severe symptoms if clinical conditions require it.^5^

A normal chest X-ray does not exclude COVID-19 pneumonia, especially in cases with mild pneumonia or in the early stage of the disease.^5,6^ It has been reported that CT cannot be used as a screening test, since the positive predictive value of thoracic CT in the diagnosis of COVID-19 is 92% high while the negative predictive value is 42% ^7^ and the absence of CT findings in the early phase of the disease should not exclude the possibility of COVID-19 disease.^2,8^ Clinical It has been reported that the combination of repetitive RT-PCR test and thoracic CT examination is beneficial in cases with suspected COVID-19.^9^

Imaging findings of viral pneumonia may overlap with non-viral infections and inflammatory conditions. Some diagnostic patterns of viral pneumonia help to make differential diagnoses in the early stages of infection, to reduce unnecessary antibiotic use, and to prevent contagion.^1^ In thorax CT in viral pneumonia; reticular opacities due to interstitial inflammation, ground-glass opacity(GGO) due to alveolar edema, patchy consolidation, localized atelectasis, peribronchovascular thickening, centrilobular nodular opacities, tree in bud pattern, interlobular septal thickening, etc. findings develop, but it is reported that diagnosis cannot be made based on imaging findings alone.^10,11^ However, detection of centrilobular nodular opacities, pleural effusion, and lymphadenopathy more frequently in non-COVID-19 viral pneumonia has been reported to help differential diagnosis.^11^

Computed tomography of the thorax is used as a common diagnosis method in the diagnosis of Coronavirus Disease 2019 (COVID-19) which causes pandemics. As in the pre-pandemic period, during the COVID-19 pandemic period, the radiological differential diagnosis of other viral agents that cause pneumonia in patients with normal immunity or in immunosuppressed patients with seasonal epidemics has gained importance in early diagnosis and isolation. Therefore, it was aimed to investigate the difference between CT imaging findings defined as characteristic in COVID-19 pneumonia and CT findings detected in pneumonia due to other viral agents previously encountered.

## Materials and Methods

### Researched Patient Population

As the COVID group, 249 COVID-19 patients aged 18 years and older, who applied to our hospital, were found to have positive Reverse Transcriptase Polymerase Chain Reaction (RT-PCR) in the nasopharyngeal swab samples taken at the application, and pneumonia was detected in the thorax CT examination at admission has been included.

For the non-COVID group, viral respiratory panel or bronchoalveolar lavage/blood viral PCR results within an average of 5.67±7.95 days, from the last 5 years before the pandemic, 18 years of age and older, with thorax CT findings compatible with viral pneumonia 94 patients who were positive but no bacterial or fungal agents were detected in other sputum and blood cultures (Viral panel results; Influenza AB n=26, Adenovirus n=5, CMV n=28, RSV n=8, Parainfluenza n=10, HMPV n=3, Endemic Coronaviruses (HCoV-NL63, HCoV-HKU, HCoV-229E, HCoV-OC43) n=16, Rhinovirus n=7, Bokavirus (HBoV) n=1) were included in the study.

### Laboratory PCR Test Method

FTD Respiratory pathogens 21 (fast-tract DIAGNOSTICS, Luxembourg) kit, which is based on the reverse transcriptase Multiplex PCR method, was used for the Viral Respiratory Panel. Artus CMV QS-RGQ kit QIAsymphony RGQ system (QIAGEN) as a CMV DNA quantitative test between January 2015 and September 2018 (measuring range of the kit: 79.4 copies / mL-100,000,000 copies / mL, 1 copy / mL = 1.64 IU / mL), COBAS Ampliprep / taqman CMV test and COBAS Ampliprep / Taqman system were used between September 2018 and December 2019 (measuring range of the kit: 150 copies / mL-10000000 copies / mL, 1 copy / mL = 0.91 IU / mL).

Viral RNA extraction from respiratory samples of patients with COVID-19 symptoms was performed manually with Bio-Speedy® Viral Nucleic Acid Isolation Kit (Bioeksen R&D Technologies Company; Turkey). RT-qPCR procedure was performed on Rotor-Gene Q 5 Plex Real Time PCR (Qiagen, Germany) using Bio-Speedy® COVID-19 RT-qPCR Detection Kit (Bioeksen Ar-Ge Technologies Company; Turkey). In the working principle of this kit, human ribonuclease P (RNA ace p) gene is targeted as an internal control. The positivity of RNAse P allows evaluation of the RT-qPCR process by confirming the extraction process, and the SARS-CoV-2 PCR result is interpreted as positive with the detection of the amplification curve of the RdRp gene region.

### Thorax CT Examination Protocol, Evaluation and Statistical Analysis

Thorax CT examination protocol; tube voltage 120kV with 64 detectors, Aquillion, Toshiba and 16 detectors Brilliance, Philips; tube current modulation 50-150 mA; range 0.85-1.4; image slice thickness is 1 mm-5 mm, CT images obtained in the supine position in full inspiratory in all patients are −600 to +1600 HU for lung parenchyma, +50 to +350 HU for mediastinum using window width; it was retrospectively analyzed using the PACS System. CT findings were evaluated by two radiologists with 5 and 20 years of experience who did not know to which group the patient belonged, and it was decided by consensus.

Age, gender, known chronic disease of the patients; CT findings include the percentage of involvement, number of lesions, distribution preference, dominant pattern, ground-glass opacity distribution pattern, nodule, tree in bud sign, interstitial changes, crazy paving sign, reversed halo sign, vacuolar sign, halo sign, vascular enlargement (vascular structures with increased calibration relative to the proximal, which is thought to be due to mediators that cause hyperemia, in the area of inflammation or in the periphery of the lesion ^12^), linear opacities, traction bronchiectasis, peribronchial wall thickness, air trapping, pleural retraction, pleural effusion, pericardial effusion, cavitation, mediastinal/hilar lymphadenopathy, dominant lesion size, consolidation, subpleural curvilinear opacities, air bronchogram, pleural thickening were examined. CT findings were also evaluated with the RSNA consensus guideline and the CORADS scoring system, data obtained were divided into two main groups as non-COVID-19 pneumonia and COVID-19 pneumonia; statistically compared with chi-square tests and multiple regression analysis of independent variables.

## Results

In the study, the age ranged between 18 and 91, with a mean of 51.99±16.99, with a median value of 53. The age of the non-COVID-19 patient group ranged from 18 to 84, with a mean of 49.29±19.43. The age of the COVID-19 patient group ranged from 18 to 91, with a mean of 53.01±15.91. In the study, 59.5% (n=204) of the patients were male, while 40.5% (n=139) were female. While 58.5% (n=55) of the non-COVID-19 pneumonia patient group were male, 41.5% (n=39) were female. Of the COVID-19 pneumonia patient group, 59.8% (n=149) were male, while 40.2% (n=100) were female. (Table 1)

**Table 1.**
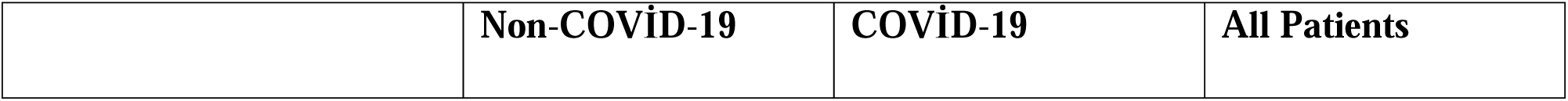
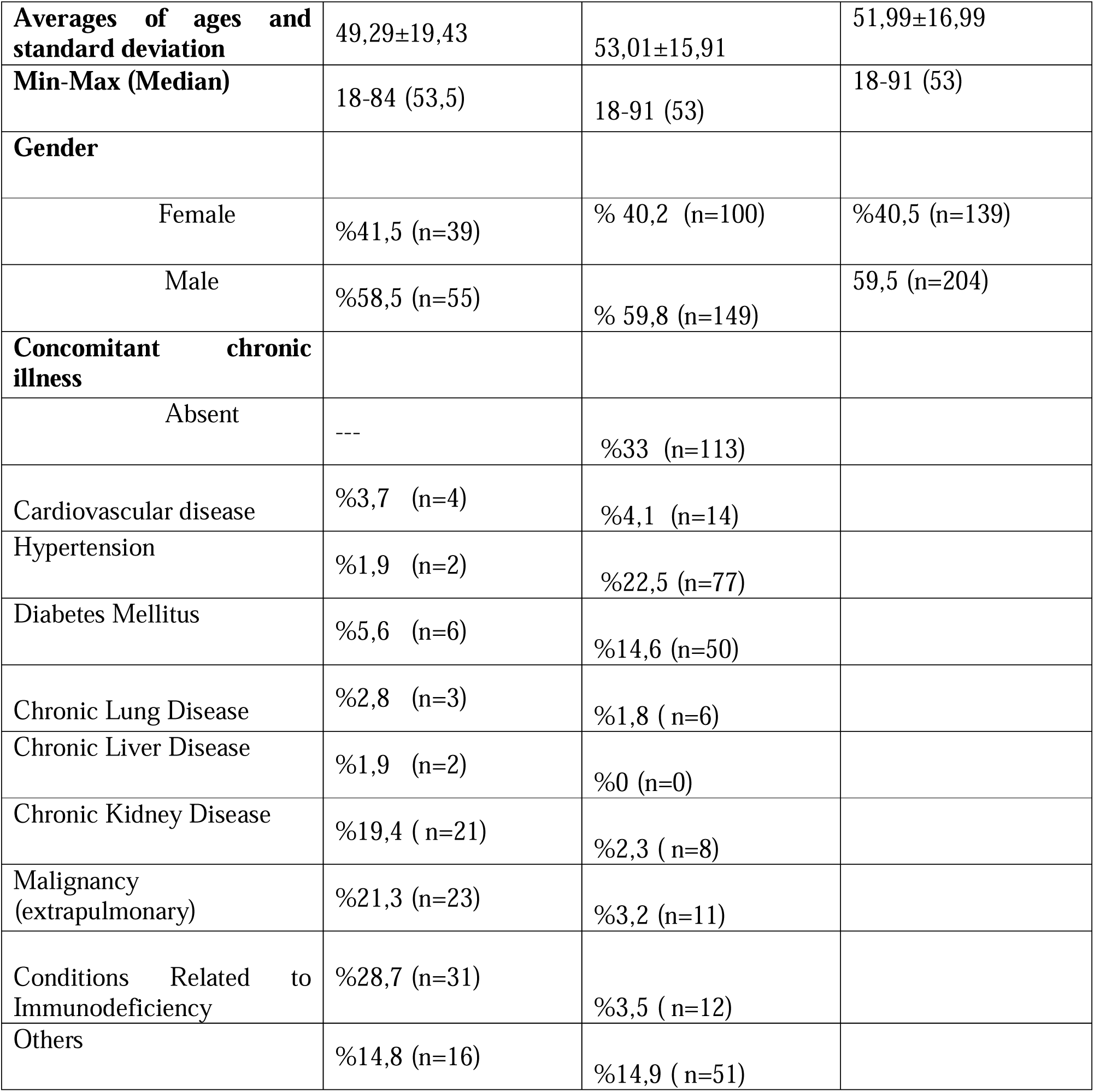

While 33% (n=113) of the COVID-19 patient group had no chronic disease, the entire non-COVID-19 patient group (n=94) had chronic diseases. Concomitant chronic diseases of COVID-19 patients including cardiovascular disease (%4,1 n=14) vs (%3,7 n=4); hypertension (%22,5 n=77) vs (%1,9 n=2); diabetes mellitus (%14,6 n=50) vs (%5,6 n=6); chronic lung disease (%1,8 n=6) vs (%2,8 n=3); chronic liver disease (%0, n=0) vs (%1,9 n=2); chronic kidney disease (%2,3 n=8) vs (%19,4 n=21); extrapulmonary malignancy (%3,2 n=11) vs (%21,3 n=23); conditions related to immunodeficiency (%3,5 n=12) vs (%28,7 n=31); others (%14,9 n=51) vs (%14,8 n=16) compared to non-COVID-19 patients. (Table 1)

In COVID-19 patients finding including RSNA typical group (%85,9 n=214) vs (%40,4 n=38); RSNA indetermine group (%11,7 n=29) vs (%34 n=32); CORADS 5 score (%77,9 n=194) vs (%26,6 n=25), CORADS 4 score (%12,4 n=31) vs (%12,8 n=12), CORADS 3 score (%6,8 n=17) vs (%31,9 n=30), CORADS 2 score (%2,8 n=7) vs (%28,7 n=27) compared to non-COVID-19 patients (p<0,01). (Table 2)

**Table 2.**

|  |  | Group |  | p |
| --- | --- | --- | --- | --- |
|  |  | Non-COVID-19 | COVID-19 |  |
| **RSNA**<br>**Consensus** | **Typical** | 38 (%40,4) | 214 (%85,9) | **0,001\*\*** |
|  | **Indetermine** | 32 (%34) | 29 (%11,7) |  |
|  | **Atypical** | 24 (%25,5) | 6 (%2,4) |  |
| **CORADS** | **CORADS 2 /Low** | 27 (%28,7) | 7 (%2,8) |  |
|  | **CORADS 3/Indetemine** | 30 (%31,9) | 17 (%6,8) |  |
|  | **CORADS 4 /High** | 12 (%12,8) | 31 (%12,4) |  |
|  | **CORADS 5 /Very high** | 25 (%26,6) | 194 (%77,9) |  |
| **Percentage of**<br>**Involvement** | **%0-%25** | 32 (%34) | 107 (%43) |  |
|  | **%25-%50** | 29 (%30,9) | 97 (%39) |  |
|  | **%50-%75** | 17 (%18,1) | 34 (%13,7) |  |
|  | **%75<** | 16 (%17) | 11 (%4,4) |  |
| **Number of**<br>**lesions** |  |  |  | **0,022\*** |
|  | **Single** | 1 (%1,1) | 17 (%6,8) |  |
|  | **Multiple** | 93 (%98,9) | 232 (%93,2) |  |
| **Distribution**<br>**Preference** |  |  |  | **0,001\*\*** |
|  | **Peripheral** | 11 (%11,7) | 101 (%40,7) |  |
|  | **Central** | 2 (%2,1) | 2 (%0,8) |  |
|  | **Peripheral+ Central** | 81 (%86,2) | 145 (%58,5) |  |
| **Dominant**<br>**Pattern** |  |  |  | **0,001\*\*** |
|  | **GGO** | 53 (%56,4) | 196 (%78,7) |  |
|  | **Consolidation** | 13 (%13,8) | 46 (%18,5) |  |
|  | **Linear, Reticular**<br>**Opacity** | 2 (%2,1) | 3 (%1,2) |  |
|  | **Nodule** | 26 (%27,7) | 4 (%1,6) |  |
|  |  | Non-COVID-19 | COVID-19 |  |
| Distribution pattern<br>of GGO | Absence (-) | 10 (%10,6) | 3 (%1,2) | **0,001\*\*** |
|  | Peripheral-bilateral | 19 (%20,2) | 140 (%56,2) |  |
|  | Round-multifocal | 20 (%21,3) | 68 (%27,3) |  |
|  | Halo Sign | 1 (%1,1) | 0 (%0) |  |
|  | Diffuse | 26 (%27,7) | 4 (%1,6) |  |
|  | Perihilar-not round | 5 (%5,3) | 3 (%1,2) |  |
|  | Single sided-Not<br>Round | 13 (%13,8) | 31 (%12,4) |  |
| Nodule |  | 56 (%60,6) | 12 (%4,8) | **0,001\*\*** |
| Tree In Bud Pattern |  | 49 (%52,1) | 8 (%3,2) | **0,001\*\*** |
| Interstitial Changes | Absent | 24 (%25,5) | 60 (%24,1) | **0,001\*\*** |
|  | Septal Thickening | 33 (%35,1) | 17 (%6,8) |  |
|  | Fine Reticular<br>Opacity | 7 (%7,4) | 72 (%28,9) |  |
|  | Septal Thickening +<br>Fine Reticular<br>Opacity | 30 (%31,9) | 100 (%40,2) |  |
| ‘Crazy Paving’<br>Pattern |  | 13 (%13,8) | 76 (%30,5) | **0,001\*\*** |
| Reversed Halo<br>(Atoll) |  | 6 (%6,4) | 109 (%43,8) | **0,001\*\*** |
| Microvascular<br>Enlargement |  | 60 (%63,8) | 207 (%83,1) | **0,001\*\*** |
| Linear Opacities |  | 71 (%75,5) | 157 (%63,1) | **0,029\*** |
| Traction<br>Bronchiectasis |  | 45 (%47,9) | 155 (%62,2) | **0,016\*** |
| Non-COVID-19 COVID-19 |  |  |  | **p** |
| **Peribronchial Wall Thickening** |  | 55 (%58,5) | 82 (%32,9) | **0,001\*\*** |
| **Air Trapping** |  | 31 (%33) | 29 (%11,6) | **0,001\*\*** |
| **Pleural Retraction** |  | 54 (%57,4) | 99 (%39,8) | **0,001\*\*** |
| **Pleural Effusion** |  | 31 (%33) | 8 (%3,2) | **0,001\*\*** |
| **Pericardial Effusion** |  | 28 (%29,8) | 9 (%3,6) | **0,001\*\*** |
| **Cavitation** |  | 3 (%3,2) | 0 (%0) | **0,001\*\*** |
| **Mediastinal-Hilar Lymph Node** | **Nonspecific** | 65 (%69,1) | 229 (%92) | **0,001\*\*** |
|  | **Pathological** | 28 (%29,8) | 20 (%8) |  |
|  | **Another Reason** | 1 (%1) | 0 (%0) |  |
| **Dominant Lesion Size** |  |  |  |  |
|  | **0-3 cm** | 43 (%45,7) | 85 (%34,1) | 0,122 |
|  | **3-5 cm** | 10 (%10,6) | 46 (%18,5) |  |
|  | **5-7 cm** | 8 (%8,5) | 30 (%12) |  |
|  | **>7 cm** | 33 (%35,1) | 88 (%35,3) |  |
| **Consolidation** |  | 52 (%55,3) | 137 (%55) | 0,960 |
| **Vacuolar Sign** |  | 11 (%11,7) | 22 (%8,8) | 0,270 |
| **Halo Sign** |  | 19 (%20,2) | 60 (%24,2) | 0,436 |
| **Subpleural Curvilinear Opacity** |  | 23 (%24,5) | 75 (%30,1) | 0,301 |
| **Air Bronchogram** |  | 23 (%24,5) | 49 (%19,8) | 0,340 |
| **Pleural Thickening** |  | 19 (%20,2) | 54 (%21,7) | 0,766 |

Chi-Square Testi \*\*p<0,01

In COVID-19 patients finding including percentage of involvement %75< (%4,4 n=11) vs (%17, n = 16), %50-%75 (%13,7 n=34) vs (%18,1 n=17), %25-%50 (%39 n=97) vs (%30,9 n=29), %0-%25 (%43 n=107) vs (%34 n=32); single lesion (%6,8 n=17) vs (%1,1 n=1); peripheral+ central distribution (%40,7 n=101) vs (%11,7 n=11); where the dominant pattern is ground glass (%78,7 n=196) vs (%56,4 n=53), consolidation (%18,5 n=46) vs (%13,8 n=13), linear-reticular opacities (%1,2 n=3) vs (%2,1 n=2), nodules (%1,6 n=4) vs (%27,7 n=26); distribution pattern of GGO absence (%1,2 n=3) vs (%10,6 n=10), peripheral-bilateral (%56,2 n=140) vs (%20,2 n=19), round-multifocal (%27,3 n=68) vs (%21,3 n=20), halo sign (%0 n=0) vs (%1,1 n=1), diffuse (%1,6 n=4) vs (%27,7 n=26), perihilar-not round (%1,2 n=3) vs (%5,3 n=5), single sided-not round (%12,4 n=31) vs (%13,8 n=13); presence of nodule (%4,8 n=12) vs (%60,6 n=56); tree in bud pattern (%3,2 n=8) vs (%52,1 n=49); interstitial changes absent (%24,1 n=60) vs (%25,5 n=24), septal thickening (%6,8 n=17) vs (%35,1 n=33), fine reticular opacity (%28,9 n=72) vs (%7,4 n=7), both septal thickening and fine reticular opacity (%40,2 n=100) vs (%31,9 n=30); crazy paving pattern (%30,5 n=76) vs (%13,8 n=13); reversed halo (%43,8 n=109) vs (%6,4 n=6); microvascular enlargement (%83,1 n=207) vs (%63,8 n=60); linear opacities (%63,1 n=157) vs (%75,5 n=71); traction bronchiectasis (%62,2 n=155) vs (%47,9 n=45); peribronchial wall thickening (%32,9 n=82) vs (%58,5 n=55); air trapping (%11,6 n=29) vs (%33 n=31); pleural retraction (%39,8 n=99) vs (%57,4 n=54); pleural effusion (%3,2 n=8) vs (%33 n=31); pericardial effusion (%3,6 n=9) vs (%29,8 n=28); cavitation (%0 n=0) vs (%3,2 n=3); mediastinal lymph node nonspecific (%92 n=229) vs (%69,1 n=65), pathological (%8 n= 20) vs (%29,8 n=28), another reason (%0 n=0) vs (%1 n=1) compared to non-COVID-19 patients (p<0,01). (Table 2)

In COVID-19 patients finding including dominant lesion size 0-3 cm (%34,1 n=85) vs (%45,7 n=43), 3-5 cm (%18,5 n=46) vs (%10,6 n=10), 5-7 cm (%12 n=30) vs (%8,5 n=8), >7 cm (%35,3 n=88) vs (%35,1 n=33); consolidation (%55 n=137) vs (%55,3 n=52); vacuolar sign (%8,8 n=22) vs (%11,7 n=11); halo sign (%24,2 n=60) vs (%20,2 n=19); subpleural curvilinear opacity (%30,1 n=75) vs (%24,5 n=23); air bronchogram (%19,8 n=49) vs (%24,5 n=23); pleural thickening (%21,7 n=54) vs (%20,2 n=19) compared to non-COVID-19 patients (p>0,05). (Table 2)

In the multiple linear regression analysis performed to determine the effect of independent variables on COVID-19 pneumonia; when the regression coefficients were examined, it was found that those with reversed halo sign (β = 0.097, p <0.05) and those with pleural effusion (β = 10.631, p <0.05) had a significant effect on COVID-19 pneumonia; it was found that COVID-19 pneumonia was more common in patients with reversed halo sign compared to those without pleural effusion. (Table 3)

**Table 3.**
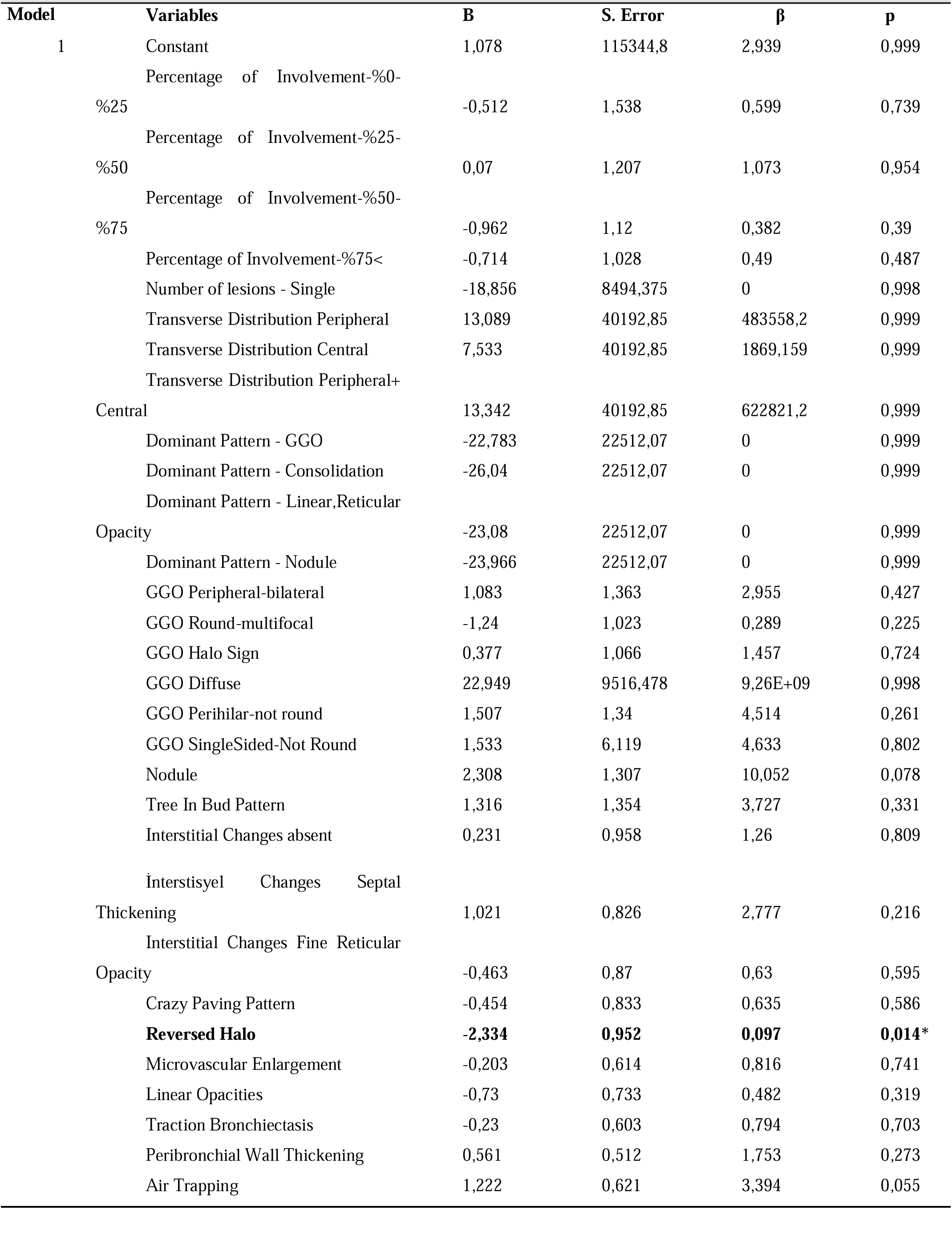
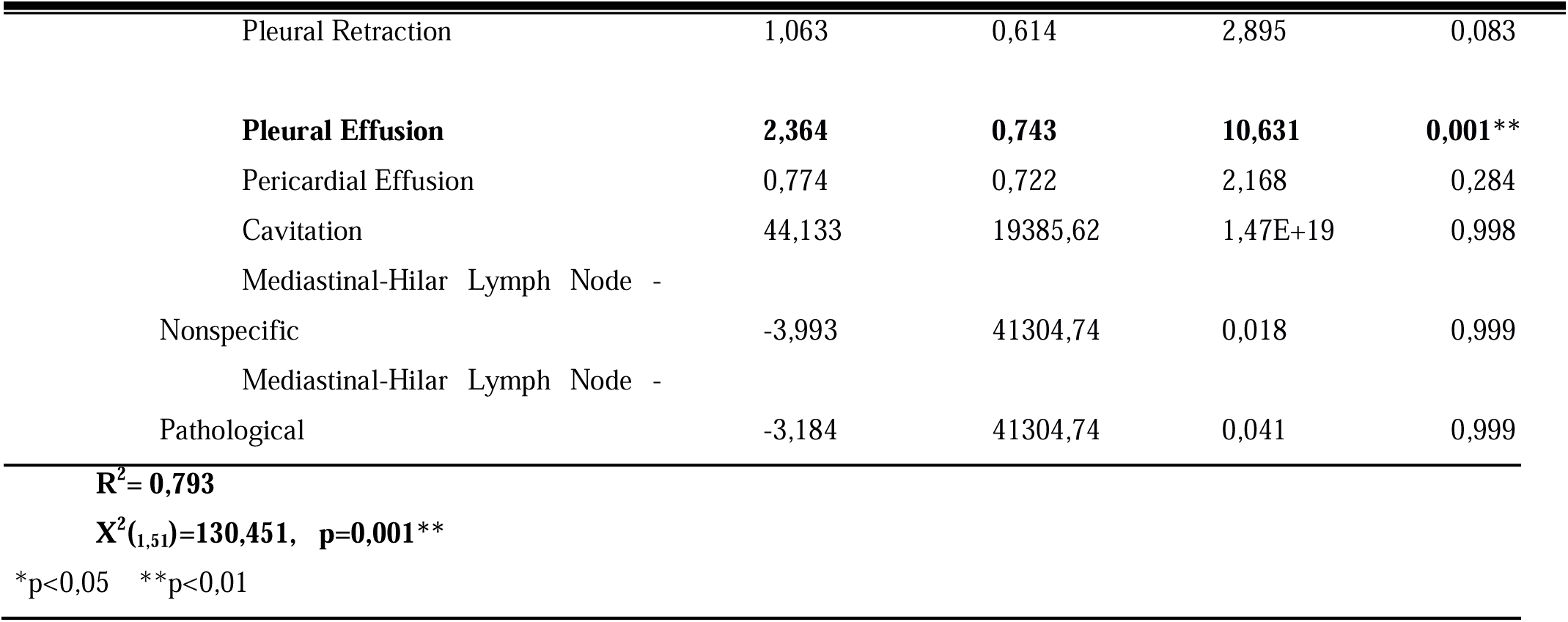
Multiple Regression Analysis Findings in Relation of Independent Variables to COVID-19

## Discussion and Conclusion

The gold standard method in the screening and diagnosis of COVID-19 is the RT-PCR test. Since the thorax CT examination is the most commonly used method in clinical practice after the RT-PCR test in the diagnosis of COVID-19, it was aimed to investigate whether the characteristic imaging findings diagnosed for COVID-19 pneumonia and whether the classification systems established for the standardization of these findings differ from the CT findings detected in pneumonia due to other viral agents.

Pleural effusion is a more common finding in non-COVID-19 viral pneumonia than in COVID-19 pneumonia^11^ (Figure 1). Although this information supports our results, in our study, all of the patients with non-COVID-19 viral pneumonia had concomitant chronic diseases, while 33% (n = 113) of the patient group with COVID-19 pneumonia had no chronic disease. The higher prevalence of diseases such as cardiovascular disease, chronic renal failure, extrapulmonary malignancy, and immunodeficiency-related conditions (73.1% vs 13.1%) in the non-COVID-19 viral pneumonia patient group also contributed to the significance of pleural effusion as a result of regression analysis may have been found.

**Figure 1.**
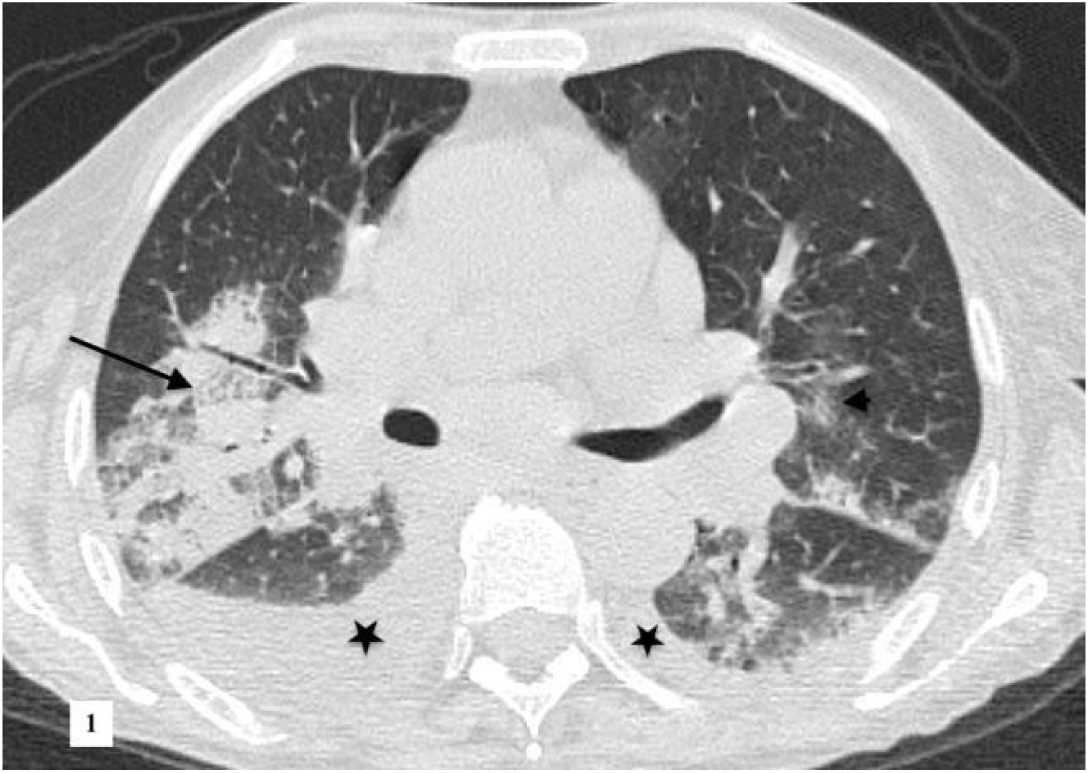
A female patient in her 70s, diagnosed with HCoV-OC43 pneumonia, chronic lymphocytic leukemia (CLL). According to the RSNA guidelines ‘typically’, the CORADS score was evaluated as 5. GGO (crazy paving) (black arrow) accompanied by interlobular and intralobular septal thickening on the axial CT section and patchy consolidation areas, faint GGO areas (black arrowhead); pleural effusion (asterisks).

While COVID-19 pneumonia often involves peripheral (Figure 2.); central or random multilobar distribution with peribronchovascular pure consolidation is observed in influenza pneumonia (Figure 3., 4., 5.). In addition, it is reported that the presence of round opacities, interlobular septal thickenings/ crazy paving, sharper lesion margin, and the absence of nodule/tree in bud appearance are helpful features for COVID-19 pneumonia to distinguish it from influenza. ^13^,^14^,^15^

**Figure 2.**
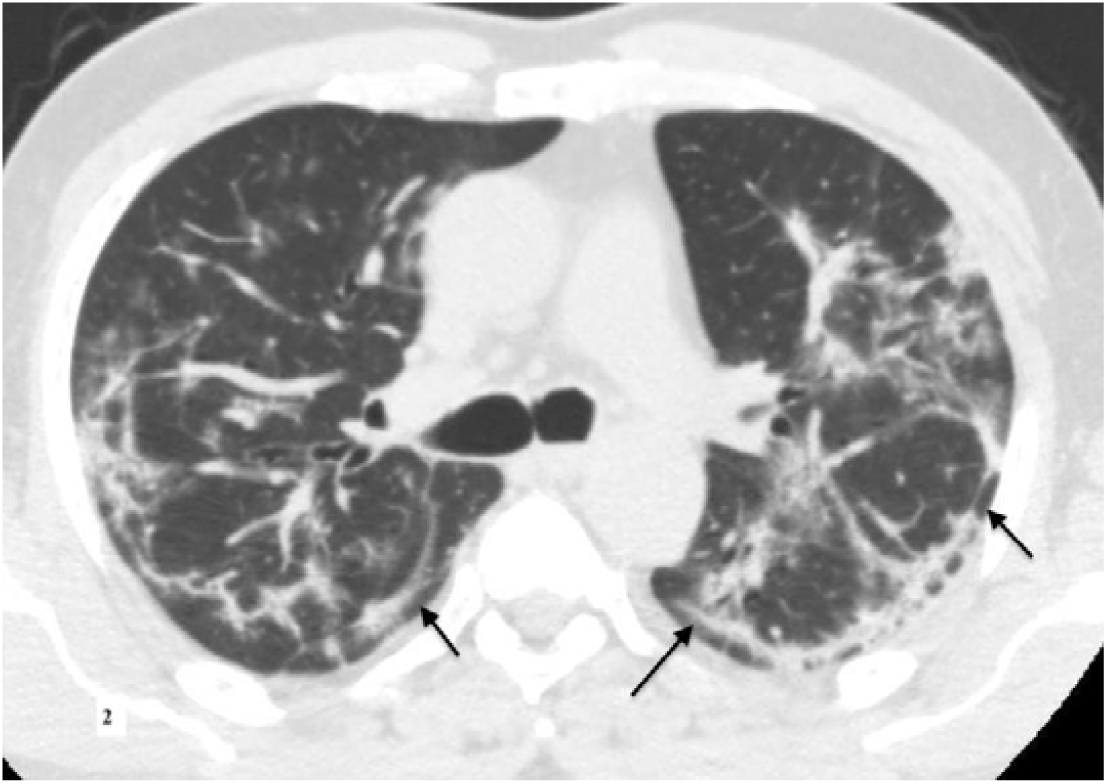
A male patient in his 50s, with COVID-19 pneumonia, known history of hypertension. ‘Typical’ according to RSNA guidelines, CORADS score was evaluated as 5. Bilateral widespread subpleural curvilinear opacities are demonstrated (black arrows).

**Figure 3.**
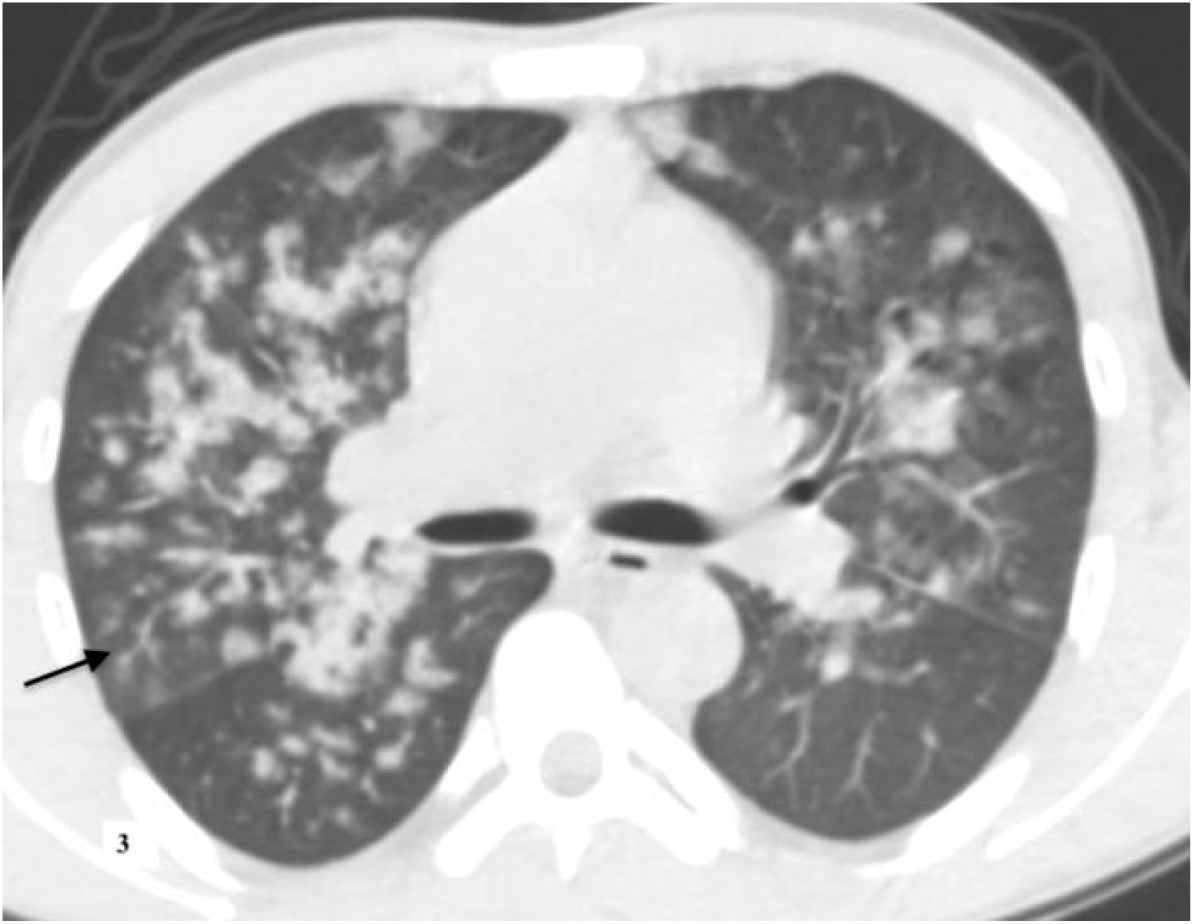
A male patient in his 30s, with influenza B pneumonia, diagnosed with known end-stage renal disease. The score was evaluated as 2 according to the CORADS classification and in the atypical group according to the RSNA guidelines. Soft tissue density centrilobular nodules (black arrow) forming tree in bud pattern and peribronchovascular consolidation.

**Figure 4.**
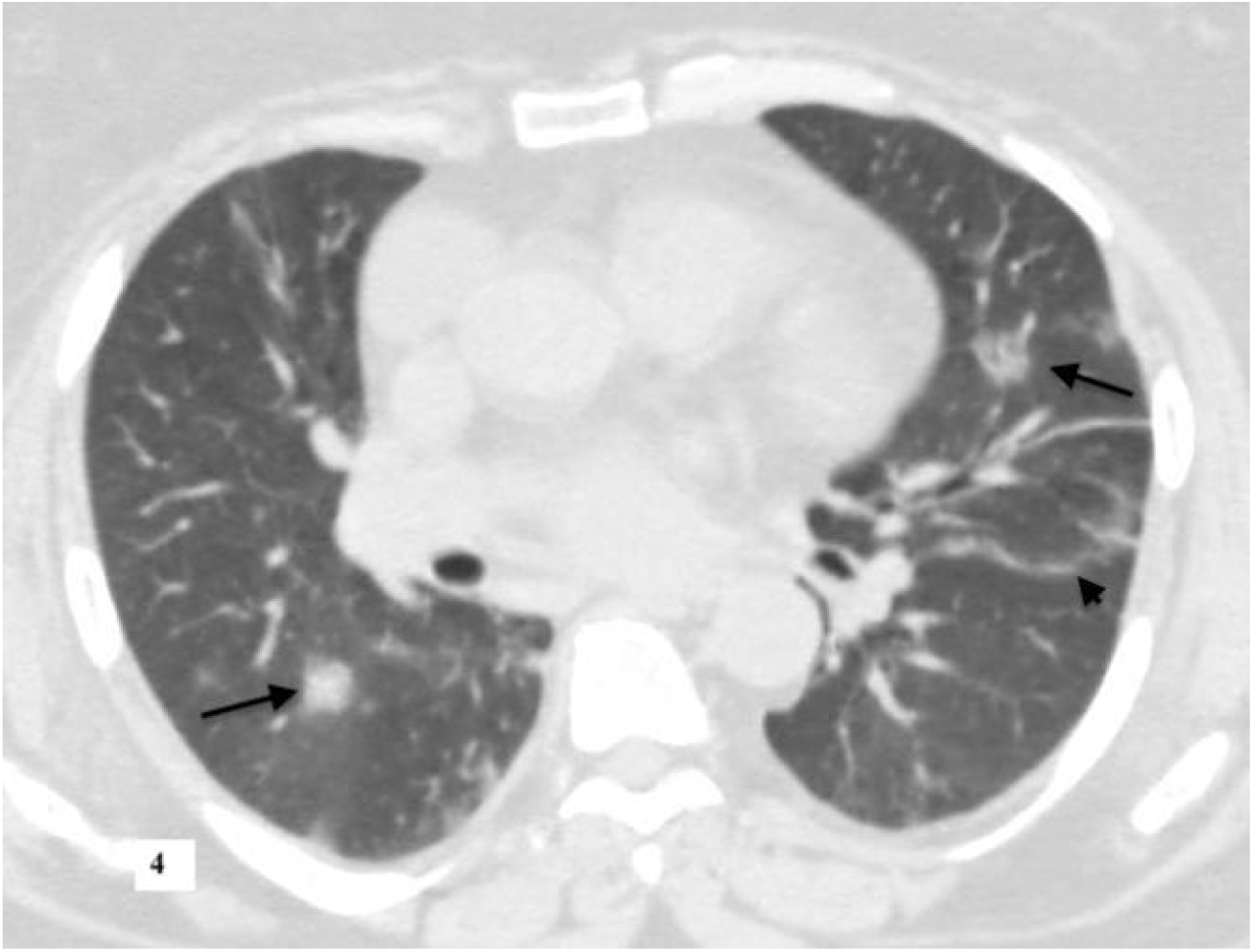
A female patient in her 60s, with influenza A (H1N1) pneumonia, known diabetes, chronic kidney disease. According to the RSNA guidelines in the typical group, the score 5 was evaluated in the CORADS classification. Bilateral rounded consolidation areas (black arrows) and parenchymal band (black arrowhead) are observed.

**Figure 5.**
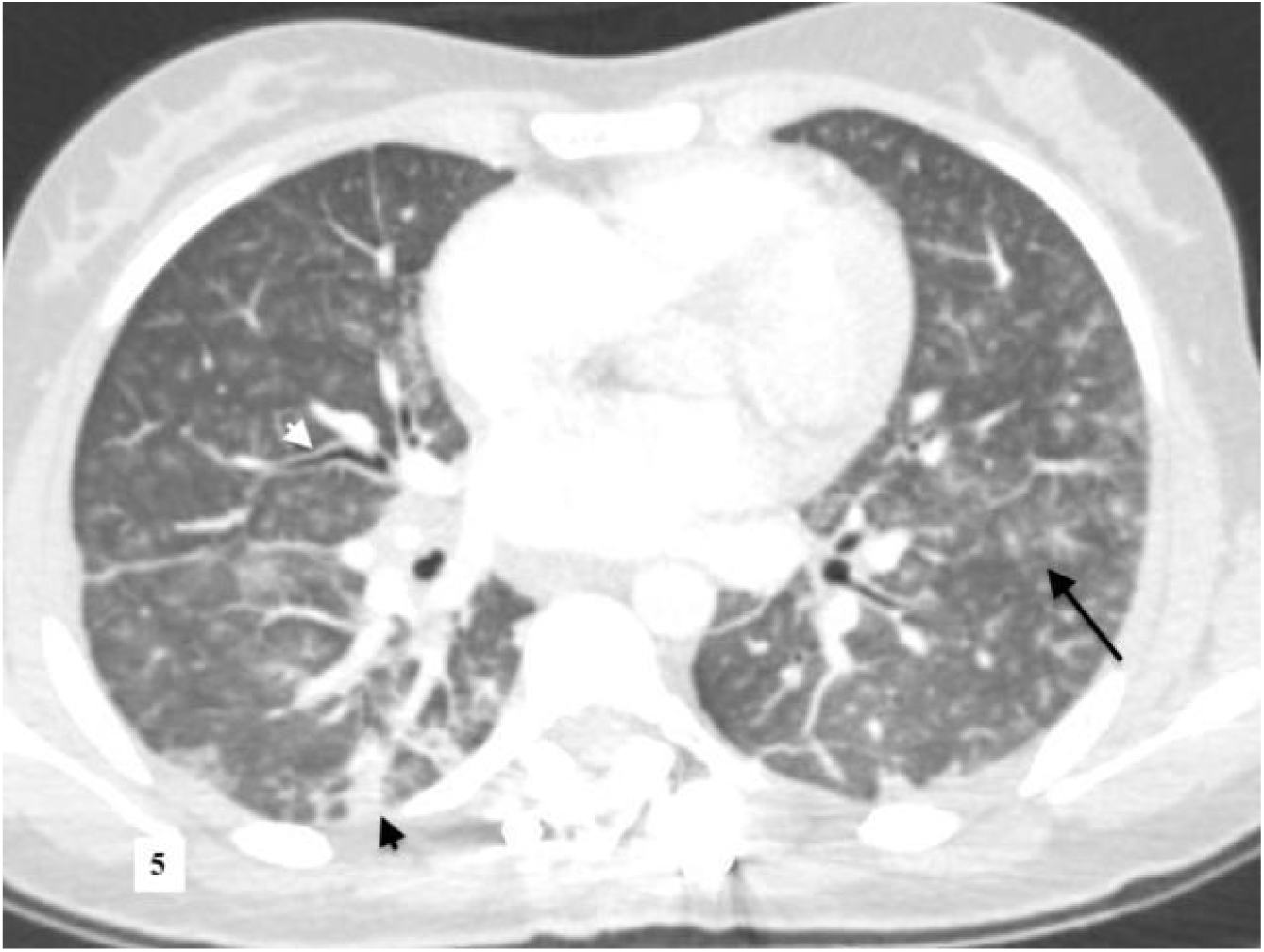
An female patient in in the late 2nd decade, with parainfluenza (HPIV 3) pneumonia, with bone marrow transplantation due to acute lymphoblastic leukemia. According to the RSNA guidelines in ‘indetermine’, CORADS score was evaluated as 3. Diffuse centrilobular ground glass density nodules (black arrow) and focal peripheral consolidation areas (black arrowhead), increased peribronchial wall thickness (white arrowhead) are observed.

In the literature, there is information that the pulmonary target sign, which is defined as a variant of the reversed halo sign by making a difference with the hyperdense dot sign in the center, is diagnostic in COVID-19 viral pneumonia.^16,17^ In our study, we did not evaluate the presence of central hyperdense dot as a separate parameter, but we think that we contributed to the literature by concluding that the presence of the reversed halo sign is valuable in differentiating other viral pneumonia from COVID-19. (Figure 6.)

**Figure 6.**
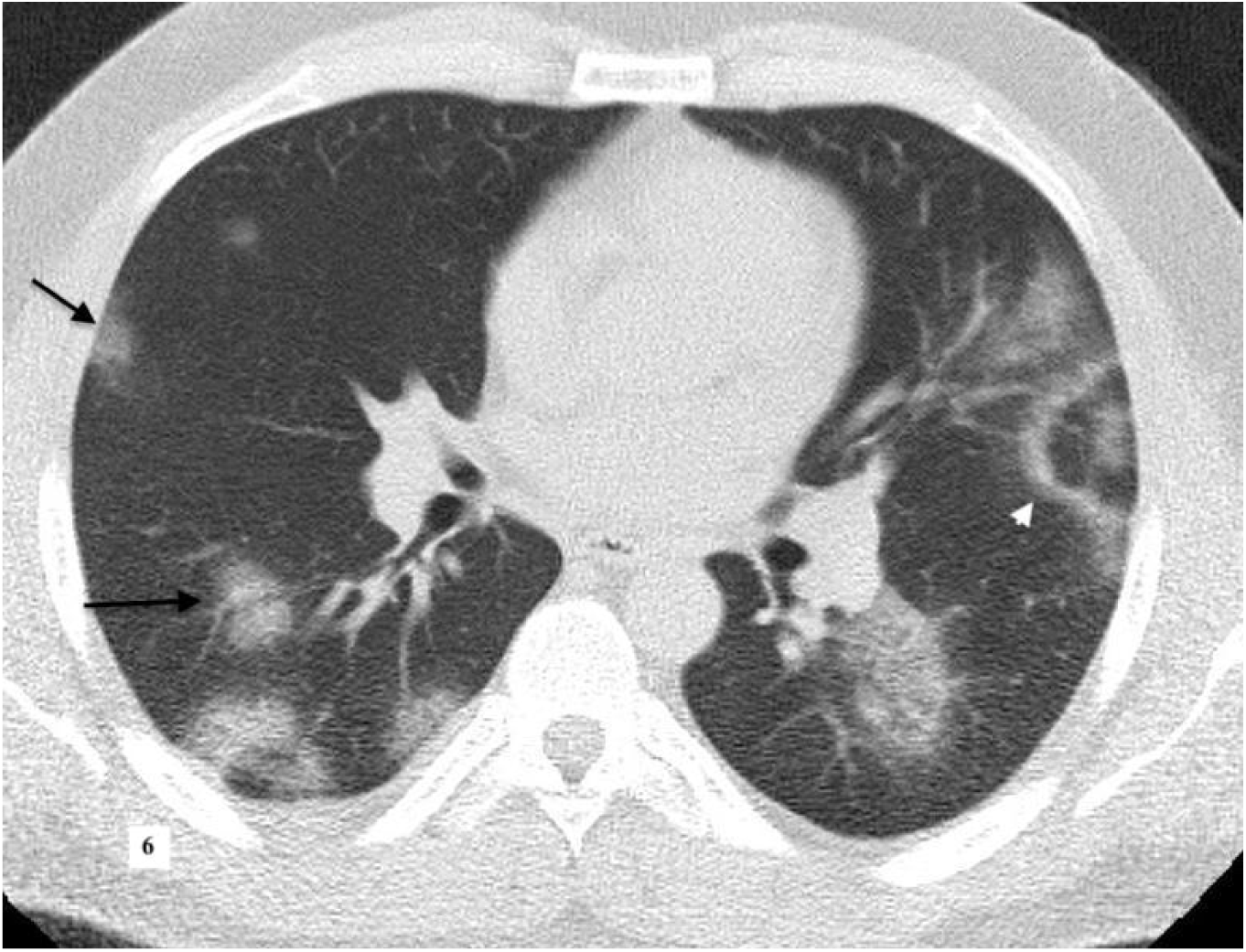
A male patient in his 30s, with COVID-19 pneumonia, known diagnosis of asthma. ‘Typical’ according to RSNA guidelines, CORADS score was evaluated as 5. Bilateral lung parenchyma rounded, multifocal GGO lesions (black arrows); reversed halo sign (white arrow) central part is relative normal, with GGO in the periphery are observed.

In the literature^18^, it has been reported that CT findings of Adenovirus pneumonia and COVID-19 pneumonia (segmental and subpleural consolidations, air bronchogram, interlobular septal thickening, accompanying mildly limited GGO and pleural effusion) overlap. (Figure 7)

**Figure 7.**
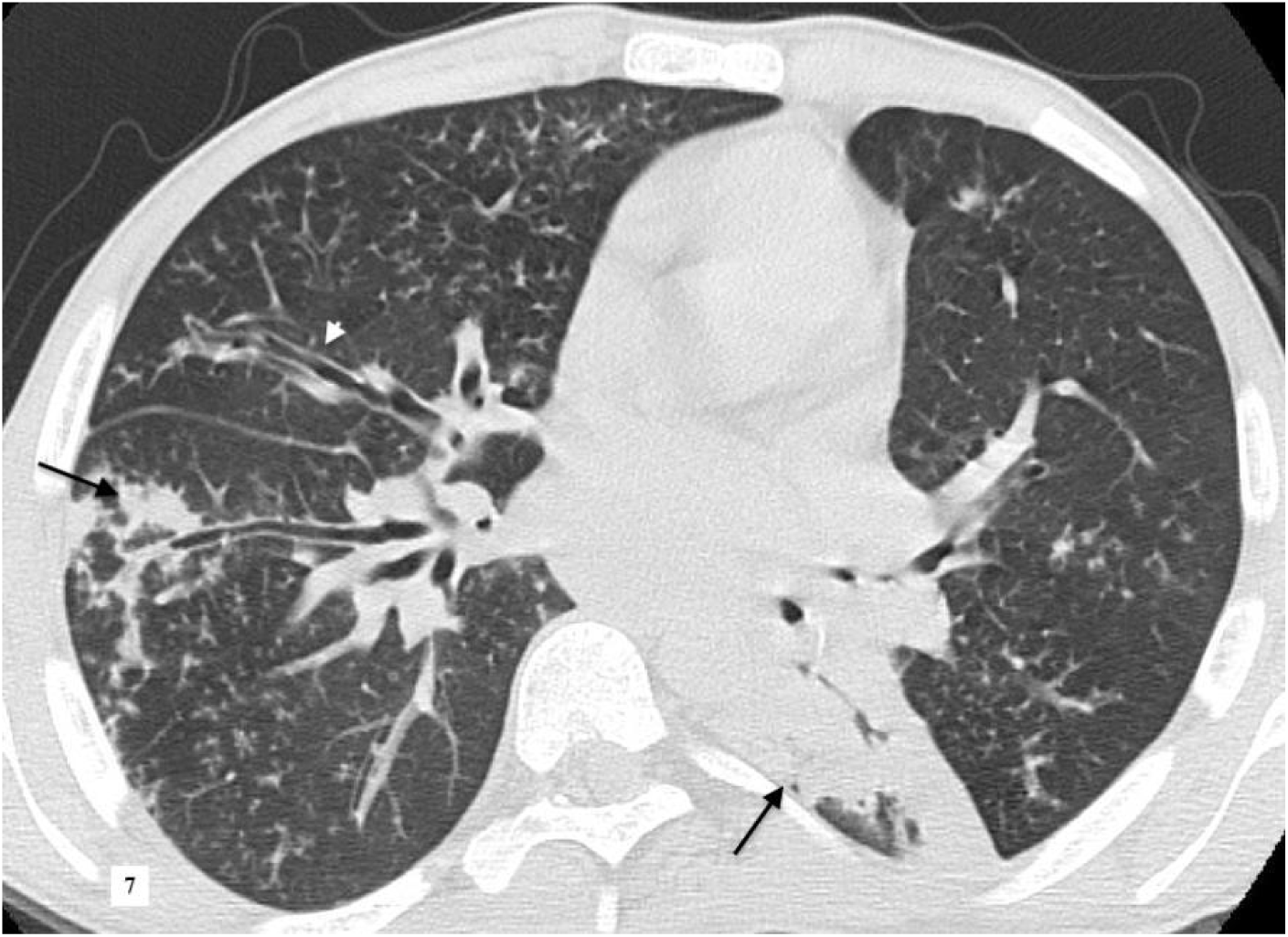
A male patient his 20s, diagnosed with known primary immunodeficiency with adenovirus pneumonia. According to the RSNA guidelines ‘indetermine’, CORADS score was evaluated as 4. İrregular peripheral consolidation (black arrows) and increased peribronchial thickness (white arrowhead) are observed.

Compared with a study ^19^ conducted with COVID-19 patients whose diagnosis was confirmed by RT-PCR and who had pneumonia on thorax CT and non-COVID-19 patients whose respiratory panel PCR was positive and pneumonia on CT, our study differed from COVID-

19. We compared two groups without COVID-19 for the RSNA consensus guideline and the CORADS system.

Classification recommendations such as RSNA consensus guideline ^2^, CORADS ^20^,^21^ have been brought to the agenda in the pandemic process with the aim of defining the imaging findings in a standard way in patients who are investigated for COVID-19 pneumonia and creating a standardized reporting language in order to provide clarity in communication with other branches for patient management by reducing uncertainty in reports. Studies evaluating the diagnostic performance of CORADS report a consistent evaluation system with high positive predictions.^8^,^22^,^23^,^24^,^25^ In the literature, there is no study comparing the findings of

COVID-19 pneumonia in terms of RSNA consensus guideline and CORADS with patients diagnosed with non-COVID-19 viral pneumonia as in our study. According to the RSNA consensus guidelines; the scores of atypical group and CORADS 2, indetermined group and CORADS 3 correspond to each other and were found to be significant in favor of non-COVID-19 viral pneumonia. The RSNA typical group and the CORADS 5 score also correspond to each other and were found to be similarly significant in favor of COVID-19 pneumonia. We think that the lack of diagnostic difference between the groups in the CORADS 4 score may be due to the fact that frequent findings in other viral pneumonia such as small but peripherally localized unilateral GGO and multifocal consolidation without other typical findings are included in this category. Although it has been reported in studies reporting that dividing the RSNA indeterminate category into 3 and 4 in the CORADS system limits the intra-observer variability^8^, since these assessment systems were developed during the pandemic process, when the prevalence of COVID-19 decreases after the pandemic is over, it is an issue that needs to be studied how much it can be applied to incidental thoracic CT findings independent of the clinic and In the future, these studies may contribute to improving the diagnostic efficiency of CORADS.

The low number of parameters affecting the result in regression analysis may be due to the low number of patients. The limited number of patients is associated with inclusion of patients with positive PCR test and proven diagnosis in the study, exclusion of cases with concomitant bacterial and fungal infections for the patient group with non-COVID-19 viral pneumonia, and the fact that patients presenting with respiratory tract infection symptoms before the COVID-19 pandemic are not performed as frequently as today, chest CT scans. The hospital information system data of the patient group diagnosed with non-COVID-19 viral pneumonia from the last 5 years before the pandemic were retrospectively scanned, and the thorax CT imaging closest to the date of PCR test was evaluated, but the CT-laboratory time interval in this group is longer than the COVID-19 pandemic period. Since the non-COVID-19 viral pneumonia group includes 5 years retrospectively, access to information about the time from the onset of symptoms to imaging is limited. The presence of co-infection in patients diagnosed with COVID-19 pneumonia is not known since most of these patients did not have additional microbial culture examinations during the pandemic period, but we expect that the presence of hospital-acquired coinfection will be lower since we have evaluated the first thorax CT examinations of these patients diagnosed with COVID-19 pneumonia.

In conclusıon; ın the diagnosis of viral pneumonia, radiological imaging, which is evaluated together with laboratory examinations, especially clinical and gold-standard RT-PCR test, has an important role in diagnosis and patient management. RSNA classification and CORADS scoring system can be used to distinguish COVID-19 pneumonia from non-COVID-19 pneumonia. The presence of reversed halo sign and absence of pleural effusion was found to be efficient in the diagnosis of COVID-19 pneumonia.

## Data Availability

All data produced in the present work are contained in the manuscript

